# Under-perception of overweight, not genetic risk, tracks weight-loss goal-setting in a direct-to-consumer genetics cohort

**DOI:** 10.64898/2026.11.16.26367268

**Authors:** Olatz Mompeo-Masachs, Stuart J. Grice, Nathan R. West

**Affiliations:** FitnessGenes Ltd, Bicester Innovation Centre, Commerce House, Telford Road, Bicester OX26 4LD, United Kingdom

**Author notes:** Correspondence: Olatz Mompeo-Masachs.

**Keywords:** weight self-perception, weight misperception, weight-loss goals, digital health, FTO, rs9939609, childhood obesity, direct-to-consumer genetics

## Abstract

Services that rely on people to recognise their own weight and set their own goals may miss those who do not see themselves as overweight. We examined how adults perceive their weight, whether genetic risk of obesity, measured body size or early weight history explains who under-perceives it, and how perception relates to the weight-management goal they set, in a direct-to-consumer genetics cohort that includes people who have not sought care for their weight (FitnessGenes; N = 20,617). Although 61% of participants were overweight or obese by BMI, only 37% described themselves as such; among those overweight or obese by BMI, 64.0% placed themselves in a lighter category than their BMI indicated. Under-perception was more common in men than in women (73.5% vs 51.6%), in younger adults and at lower BMI, and a sex difference in the same direction, though smaller, was present when non-recognition of overweight was assessed against waist-to-height ratio rather than BMI. FTO rs9939609 behaved as an obesity risk variant (+0.46 kg/m² BMI and OR 1.18 for recalled childhood obesity per risk allele) but did not predict under-perception at a given BMI (per-allele OR 0.97, 95% CI 0.92–1.03), and neither did MC4R rs17782313 nor a combined score. A recalled history of childhood obesity, by contrast, was associated with heavier self-perception at the same adult BMI (+10.0 percentage points). At the same BMI, participants who under-perceived their weight were about half as likely to set a weight-loss goal (26.8% vs 47.5%; OR 0.52, 95% CI 0.48–0.56); among those obese by BMI, 49% who recognised it chose weight loss, against 18% who did not. Genotype did not predict goal- setting beyond its effect on BMI. Men were less likely both to recognise overweight and, having recognised it, to choose weight loss. Genetic risk of obesity therefore identifies who is likely to be heavier but not who will recognise it, and self-perception marks a group, mainly men and those at lower BMI, whom self-directed weight-management services are likely to under-reach.

## 1 Introduction

Direct-to-consumer (DTC) genetic testing has produced large cohorts in which genotype, body size and self-reported health behaviour are recorded for the same individuals. Unlike clinical samples, these cohorts include people who have not sought care for their weight and may not regard it as a concern. They therefore show how people judge their own health, and how that judgement relates to what they intend to do about it, at a scale clinical datasets rarely reach.

Whether a person acts on their weight depends in part on recognition: whether they perceive themselves as overweight. Many adults who are overweight by clinical criteria do not describe themselves as such, and those who do not recognise their overweight are less likely to attempt weight management (Duncan et al., 2011; Gruszka et al., 2022). The size of the shortfall varies with setting, sex and the measure used: among 540 adult women in Erbil, Iraq, 79.1% classified their weight status correctly and only 13.3% underestimated it, although underestimation still outnumbered overestimation (Shabu et al., 2024). As overweight has become more prevalent, the reference against which people judge their own size appears to have shifted upward with it (Robinson, 2017). Self-perception also influences whether people engage with structured weight-management programmes, including digital ones (Sharpe et al., 2017). If perception rather than measured size determines intention, services that depend on self-referral or self-set goals will reach people according to how they see themselves rather than according to need. Who under-perceives, and whether under-perception relates to the goals people set, are therefore practical questions for such services.

Under-perception may also have determinants beyond current body size. One candidate is genetic. Body weight is partly heritable, and common genetic variants confer measurable risk of obesity (Loos and Yeo, 2022). The most robustly replicated of these lies at the FTO locus: each copy of the rs9939609 A allele is associated with higher body weight and adiposity, across ancestries and from childhood onwards (Frayling et al., 2007; Yang et al., 2012; Loos and Yeo, 2022), largely through increased appetite and energy intake (Cecil et al., 2008; Speakman, 2015). FTO has also been linked to eating behaviour and affect beyond its effect on BMI (Harbron et al., 2014; Koike et al., 2018), and in an exploratory study of thirty healthy young men the risk allele was associated with higher emotional eating scores (Brown et al., 2026). None of this work examined body perception directly, but it makes an influence on how carriers appraise their own bodies plausible enough to test. Some FTO associations, such as those with type 2 diabetes and related metabolic traits, are explained entirely by its effect on adiposity (Frayling et al., 2007; Freathy et al., 2008); others, such as that with depression, persist after adjustment for BMI (Samaan et al., 2013). Whether genetic risk of obesity shapes self-perception independently of body size has not, to our knowledge, been tested. If it did, genotype could help identify people likely to under- perceive their weight; if it does not, genetic risk information alone cannot.

A second candidate is developmental. Higher childhood BMI prospectively predicts later body dissatisfaction (Blundell et al., 2024), and an early history of obesity may set the reference against which adults judge their own size.

We used a DTC genetics cohort in which genotype, self-reported height and weight, self-perceived body type, recalled childhood obesity and a stated weight-management goal were recorded for the same individuals. We asked four questions. First, how closely self-perceived body type agrees with BMI category, and who under-perceives. Second, whether genetic risk of obesity, at FTO rs9939609 and independently MC4R rs17782313, is associated with under-perception at a given BMI, having first confirmed that these variants confer the expected higher BMI and childhood obesity in this cohort.

Third, whether a recalled history of childhood obesity is associated with under-perception at a given BMI. Fourth, whether under-perception is associated with the weight-management goal a person sets, and whether genotype adds to measured size in predicting it.

## 2 Materials and Methods

### 2.1 Study design and participants

We performed a cross-sectional analysis of the FitnessGenes direct-to-consumer genetics cohort. Participants were adult customers at registration who had genetic testing and completed an onboarding questionnaire capturing self-reported anthropometry, self-perceived body type, and childhood obesity history. Association models required non-missing FTO rs9939609 risk and self-perceived body type (N = 20,695). Analyses that additionally required a BMI category — the sample description in Table 1 and the concordance analyses of Section 3.3 — were restricted to the 20,617 participants with all three (BMI category was missing for 78 otherwise-eligible participants). Thirty-two records (0.15%) carry an implausible recorded age — 21 below 18 years and 11 above 100, including five near 950 years, consistent with placeholder birth dates. Age enters the models only as a covariate and excluding these records changes no estimate materially (for example, the per-allele FTO association with self-perception moves from +2.43 to +2.39 percentage points), so they were retained; Table 1 reports the age distribution both with and without them. Every analytic sample used in the paper is listed with its inclusion criteria in Supplementary Table S3. The mediation sample additionally required recalled childhood obesity status (N = 10,637; the high-versus-low-risk contrast used for mediation comprised 5,626). Registration and questionnaire dates were not retained in the de-identified extract, so the accrual window cannot be specified. Customer data were used for research under the consent recorded at registration.

**Table 1.** Characteristics of the primary analytic sample (N = 20,617).

| Characteristic | Value |
|---|---|
| Age, mean (SD), years | 45.0 (12.5), range 18–93 y, excluding 32 implausible values; 45.3 (19.1) with them retained |
| Female, % | 48.1 |
| BMI, mean (SD), kg/m<sup>2</sup> | 27.7 (6.5) |
| BMI category: normal, % | 37.6 |
| BMI category: overweight, % | 34.8 |
| BMI category: obese I–III, % | 26.4 |
| BMI category: underweight, % | 1.2 |
| Self-perceived: underweight, % | 6.7 |
| Self-perceived: lean/fit, % | 21.7 |
| Self-perceived: average, % | 34.4 |
| Self-perceived: overweight, % | 27.7 |
| Self-perceived: obese, % | 9.4 |
| FTO rs9939609: TT / TA / AA, % (risk allele A) | 37.5 / 46.8 / 15.7 |
| MC4R rs17782313: AA / AG / GG, % (risk allele G) | 58.0 / 35.9 / 6.1 |
| Genotyping source: array / uploaded, % | 67.9 / 32.1 |
| Ethnicity of those reporting: white, % | 76.6 |
| Ethnicity of those reporting: Asian, % | 7.5 |
| Ethnicity of those reporting: other, Hispanic, Black or mixed, % | 15.9 |

*Ethnicity was missing for 44.1% of participants. MC4R rs17782313 percentages are among the 20,117 participants with a called genotype (2.4% uncalled in this sample; 3.9% across the full extract, which contains a larger share of uploaded genotypes); a called FTO genotype is an inclusion criterion, so FTO has no such missingness.*

### 2.2 Ethics statement

The study was reviewed by the FitnessGenes internal Ethics Committee (reference FGEC/2026/012), which confirmed valid opt-in research consent, compliance with UK GDPR, and anonymisation of the dataset. The study was conducted in accordance with the Declaration of Helsinki and the local legislation and institutional requirements. Participants provided separate, opt-in electronic informed consent for the research use of their data at registration, before genetic testing, and could withdraw consent at any time. The research team had no access to personal identifying information.

### 2.3 Measures

**Genotyping.** FTO rs9939609 genotype was recorded as TT / TA / AA and mapped to a three-level risk grouping (TT = low, TA = moderate, AA = high; risk allele = A); records with an absent or ambiguous call (n = 364, of which 15 were ambiguous) were treated as missing throughout. The primary contrast was high (AA) versus low (TT) risk; a per-risk-allele (0/1/2) coding was used for dose-response and sex- stratified models. DNA was obtained from saliva collected by the Oragene self-collection kit (DNA Genotek Inc., Ottawa, ON, Canada) and returned by post, and extracted by LGC Genomics (Hertfordshire, UK), following the sample-collection and extraction procedure described previously for a separate, smaller sample of the same provider’s customers (West et al., 2018). Genotypes in the present extract came from two sources. For 67.9% of the analytic sample, they were called from a genome-wide Illumina Infinium Global Screening Array (versions 1, 2 and 3); for the remaining 32.1%, they were taken from genotype files that customers had uploaded from third-party consumer testing services. Call rates for FTO rs9939609 were 99.9% in the array subset and 95.1% in the uploaded subset, and for MC4R rs17782313 99.8% and 88.5%, respectively (Supplementary Table S4). Uploaded genotypes were accepted as supplied and were not recalled from raw intensity data, so their platform and quality- control provenance cannot be verified. Genotypes were tested for Hardy–Weinberg equilibrium by goodness-of-fit χ² overall and by genotyping source. FTO rs9939609 is the primary exposure. MC4R rs17782313 is analysed separately as an independent check, because the two variants act through partly different biology. Its own association with BMI in this cohort is weak, however, so a matching null for MC4R is supportive rather than a true internal replication. A two-variant risk-allele score (range 0–4) is secondary, computed both unweighted and weighted by the per-allele BMI effects estimated in this cohort. Each variant and score is reported for every key outcome in Supplementary Table S2, with genotype frequencies, call rates and Hardy–Weinberg tests by genotyping source in Supplementary Table S4. The extract retained only the selected variants rather than genome-wide data, so a BMI polygenic score could not be constructed.

**Anthropometry and BMI category.** Height and weight were self-reported. BMI was calculated as weight in kilograms divided by height in metres squared and categorised using standard World Health Organization thresholds (World Health Organization, 2000): underweight (< 18.5 kg/m²), normal weight (18.5–24.9 kg/m²), overweight (25.0–29.9 kg/m²), obese class I (30.0–34.9 kg/m²), obese class II (35.0– 39.9 kg/m²) and obese class III (≥ 40.0 kg/m²). Where a binary or three-level classification was required the three obese classes were combined, and “overweight or obese by BMI” denotes BMI ≥ 25.0 kg/m². BMI was used throughout as the reference against which self-perception was judged because it was available for all participants, whereas waist measurements were recorded for about one in five; waist- to-height ratio, a better marker of central adiposity (Ashwell et al., 2012; Gibson and Ashwell, 2020), served as a sensitivity check. In a subset, waist circumference was self-reported in centimetres, and waist-to-height ratio (WHtR) was calculated as waist circumference divided by height. Physical activity was self-reported on a single five-level ordinal item (“How active you are, ranging from low to very active”) with the response options none, light, moderate, active and very active; in the primary analytic sample (N = 20,617) these were chosen by 385, 3,183, 6,410, 7,513 and 3,126 participants respectively, and the item was modelled as an equally spaced ordinal score from 0 (none) to 4 (very active).

**Self-perceived body type.** Participants selected one of five ordered categories: underweight, lean/fit, average, overweight, or obese. A binary overweight/obese self-perception (overweight or obese vs. others) was the outcome for the genotype-association models; under-classification, defined below, was the outcome for analyses of who under-perceives and of goal-setting. Two distinct quantities are reported and are not interchangeable. Under-classification denotes perceiving oneself in a lighter category than one’s BMI category, so a participant with class II obesity who describes themselves as overweight is under-classifying; failure to recognise overweight denotes not describing oneself as overweight or obese at all. Among participants who were overweight or obese by BMI, 64.0% under- classified while 41.5% did not describe themselves as overweight or obese. Each quantity is named explicitly wherever it is used. For the descriptive concordance analysis, perceived categories were mapped onto clinical BMI bins (primary mapping: lean/fit and average → normal); a more tolerant mapping, in which “average” was treated as concordant with either normal or overweight BMI, was reported as a sensitivity analysis.

**Recalled childhood obesity.** A self-reported binary indicator of childhood obesity (yes/no), retrospectively recalled and available for 10,827 participants overall, of whom 10,609 fall in the primary analytic sample. The item asked whether the participant was obese during childhood, defined in the questionnaire as before the age of 16 years, with response options yes and no. Because the item records whether obesity was present in childhood rather than an age at onset, “recalled childhood obesity” here denotes a history of childhood-onset obesity.

Weight-management goal. At onboarding, participants selected one of four goals: get fit, lose weight, gain lean muscle, or body maintenance. A binary weight-loss goal (lose weight versus any other goal) was the outcome for the analyses in Section 3.6. The item was recorded for 19,221 participants.

### 2.4 Statistical analysis

Sample characteristics were summarised as mean (SD) or percentage. We described concordance between self-perceived body type and BMI category using row-normalised cross-tabulation and classified it as concordant, under-classifying, or over-classifying. Associations between FTO risk and binary overweight/obese self-perception were estimated with linear-probability (ordinary least squares) models, giving effects directly in percentage points. Primary models were adjusted for age and sex.

These covariates are conventional rather than necessary here: genotype is fixed at conception and so is not confounded by lifestyle, and the per-allele FTO estimate is the same with and without them (+2.42 percentage points unadjusted, +2.43 adjusted). Population stratification does remain a potential confounder, and is addressed below. Models additionally adjusted for genotyping source were also examined. Heteroskedasticity-robust (HC3) standard errors were used for all linear-probability models. As a confounding sensitivity analysis, we additionally adjusted for self-reported ethnicity in the subset with ethnicity recorded. As a secondary analysis addressing a distinct question, we adjusted for BMI to ask whether any association persists independently of actual body size; because BMI lies on the causal pathway, this model is not part of the primary inference, and conditioning on a mediator can introduce collider bias if unmeasured factors influence both BMI and self-perception (VanderWeele, 2016).

Mediation of the FTO–self-perception association by recalled childhood obesity was estimated with linear-probability models: the total effect (c) from a model of the outcome on exposure; the direct effect (c′) and mediator→outcome path (b) from a model additionally including the mediator; and the exposure→mediator path (a) from a linear-probability model of the mediator on exposure. The indirect effect was estimated as a × b, which in linear models equals c − c′, and the proportion mediated as indirect/total (Preacher and Hayes, 2008; VanderWeele, 2016). Indirect effects and the proportion mediated were bootstrapped with 3,000 resamples (percentile 95% confidence intervals); the mediation decomposition is reported in Supplementary Figure S1 as a secondary analysis. To characterise the mediator, the association of per-allele FTO risk with recalled childhood obesity was also estimated by logistic regression, and the association of childhood onset with adult BMI by linear regression adjusted for age and sex. As a positive control, the per-allele association of FTO rs9939609 with BMI itself was estimated (+0.46 kg/m², 95% CI 0.33–0.59, p = 6 × 10⁻¹², n = 20,616), confirming that the genotype data behave as expected in this cohort. Sex-stratified per-allele effects and a sex-by-genotype interaction were tested, and an exploratory FTO × physical-activity interaction was examined. In a further sensitivity analysis we substituted waist-to-height ratio (WHtR) for BMI, in the subset with waist measurements. As additional robustness checks we refitted the primary association with logistic regression, restricted it to the largest ancestry group, and substituted an aggregate FTO+MC4R risk score for the single variant.

Secondary and sensitivity analyses were not corrected for multiple testing and are interpreted as supportive or exploratory. Associations with weight-loss goal-setting (Section 3.6) were estimated with the same linear-probability approach, entering self-perceived body type and BMI together so that each is estimated holding the other constant; a multinomial logistic model across all four goals with ‘get fit’ as the reference category, an ordered five-level perception predictor, restriction to the largest ancestry group, and adjustment for physical activity were examined as sensitivity analyses. Under-perception was additionally assessed against central adiposity (waist-to-height ratio ≥ 0.5) in the subset with waist measurements, and under-classification was recomputed excluding participants who described themselves as lean/fit, because that option describes fitness as well as size. Because self-perceived body type and the stated goal were reported in the same questionnaire, these associations are interpreted as recognition tracking intention rather than causing it; the composition of the four goal categories across perceived body types was examined to assess how far the association is definitional. Analyses were performed in Python 3.11.17 using pandas 2.3.3, numpy 2.4.6, scipy 1.17.1, statsmodels 0.15.0 and matplotlib 3.11.2. The study is reported in accordance with the STROBE guideline for observational studies, with the STREGA extensions for the genetic analyses.

## 3 Results

### 3.1 Sample characteristics

The primary analytic sample comprised 20,617 adults (Table 1). Mean age was 45.0 years (SD 12.5, excluding 32 implausible recorded ages; Section 2.1), 48.1% were women, and mean BMI was 27.7 kg/m² (SD 6.5); 61.2% were overweight or obese by BMI. FTO rs9939609 genotype frequencies were TT 37.5%, TA 46.8% and AA 15.7% (risk-allele frequency 0.391). Across the 20,929 records with a called genotype, genotype proportions departed mildly from those expected under Hardy–Weinberg equilibrium (p = 0.009). The departure was detected in the array-genotyped subset (p = 0.011, n = 14,265) but not in the smaller uploaded-data subset (p = 0.36, n = 6,664), which has less power to detect it; the two subsets therefore cannot be said to differ. Population structure in a multi-ancestry cohort and a genotyping artefact in the array subset both remain possible. Strand misalignment, to which this A/T variant is vulnerable, does not: the A-allele frequency is 0.391 in both subsets, whereas a systematic flip in the uploaded data would give approximately 0.61 (Supplementary Table S4). Equal allele frequencies do not, however, exclude miscalling of heterozygotes, which would also produce the observed heterozygote deficit. Recalled childhood obesity was available for 10,609 participants in the primary analytic sample (yes, 1,473; no, 9,136); models of this item that do not require a called genotype include up to 10,797 participants.

Participants with and without recalled childhood-obesity data had similar BMI (27.5 vs 27.9 kg/m²; standardised difference −0.07) and similar rates of overweight/obese self-perception (36.9% vs 37.4%; −0.01). They differed in questionnaire completeness. Throughout, completing the extended questionnaire module means having answered the childhood-obesity item. The item belongs to a module that also carried the waist, ethnicity, diabetes, family-history and emotional-eating questions: 98.8% of participants who answered it also reported ethnicity, against 10.5% of those who did not, and waist measurements were recorded only within the module. Participants who answered it also stated a weight-loss goal more often (32.3% vs 14.0%). Full comparisons are given in Supplementary Table S1.

### 3.2 Genetic risk of obesity confers higher BMI and childhood obesity

FTO rs9939609 behaved as an obesity risk variant in this cohort. Each additional A allele was associated with a 0.46 kg/m² higher BMI (95% CI 0.33–0.59, p = 6 × 10⁻¹², n = 20,616). The effect was approximately twice as large in women (+0.62 kg/m² per allele, 95% CI 0.41 to 0.82) as in men (+0.30, 95% CI 0.14 to 0.47; sex-by-genotype interaction +0.31 kg/m² per allele, 95% CI 0.04 to 0.57, p = 0.023). MC4R rs17782313 was only weakly associated with BMI (+0.14 kg/m² per allele, 95% CI −0.01 to +0.29, p = 0.07).

Associations of both variants with waist-to-height ratio follow the same pattern and attenuate fully on adjustment for BMI, indicating no effect on fat distribution independent of overall adiposity; they are reported in Supplementary Table S5.

Genetic risk was also associated with obesity in childhood. Each additional FTO risk allele increased the odds of recalled childhood obesity (OR 1.16, 95% CI 1.07–1.26, p = 2 × 10⁻⁴; adjusted for age and sex, OR 1.18, 95% CI 1.09–1.28, p = 3 × 10⁻⁵). MC4R showed an association of similar size (OR 1.16 per allele, p = 0.001), as did the combined FTO+MC4R risk-allele score (OR 1.17 per allele, 95% CI 1.10–1.24, p = 4 × 10⁻⁷). Childhood obesity in turn marked a heavier adult trajectory. Adults who reported it had a higher BMI (mean 31.6 vs 26.8 kg/m²; +4.7 kg/m² adjusted for age and sex; +5.8 in women and +3.9 in men), a higher prevalence of severe obesity (BMI ≥ 40 kg/m²: 13.3% vs 2.6%) and greater central adiposity (waist-to-height ratio +0.053, p = 8 × 10⁻³⁷; mean 0.545 vs 0.495; n = 4,271). Genetic risk therefore carried through to body size in childhood and adulthood.

### 3.3 Self-perception follows BMI but systematically understates it

Self-perceived body type rose steadily with measured BMI (Spearman ρ = 0.72), but from the ‘average’ label upward each category corresponded to a BMI about one WHO band heavier than the label implies. Mean BMI was 21.3, 24.6, 25.4, 30.7 and 38.8 kg/m² across the five perceived categories from underweight to obese, so participants who described themselves as ‘average’ were on average already in the overweight band, and those who described themselves as ‘overweight’ were on average in obese class I. Although 61% of the sample were overweight or obese by BMI, only 37% described themselves as overweight or obese; this comparison does not depend on how perceived categories are matched to BMI categories. When each perceived category was mapped to a clinical BMI category, 53.2% of participants were concordant, 44.8% placed themselves in a lighter category than their BMI indicated (under-classification) and 2.0% placed themselves in a heavier one (over-classification) (Figure 1).

**Figure 1.**
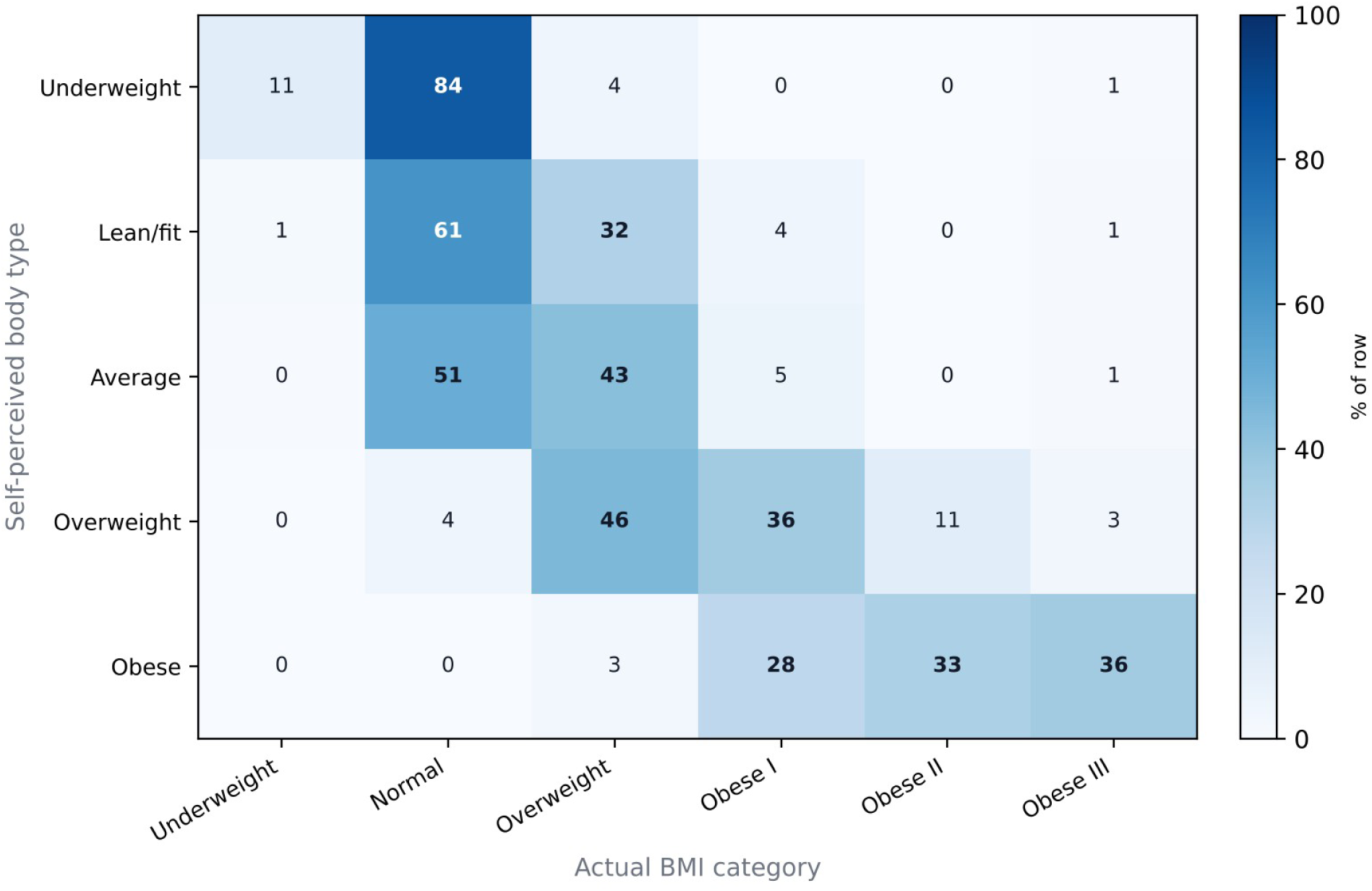
Self-perceived body type (rows) versus actual BMI category (columns), row-normalised percentages (N = 20,617).

Concordance depended on how the ambiguous ‘average’ label was mapped: treating ‘average’ as concordant with either normal or overweight BMI raised concordance to 67.8% and reduced under- classification to 30.2%, with over-classification unchanged at 2.0%. Under either mapping, misperception was almost entirely towards under-classification. Of participants who described themselves as ‘lean/fit’, 38% were overweight or obese by BMI, as were 49% of those who described themselves as ‘average’ (Figure 1).

### 3.4 Who under-perceives their weight

Under-perception was concentrated in men, younger adults and those at lower BMI within the overweight and obese range. Of the 12,840 participants who were overweight or obese by BMI and reported a body type, 64.0% under-classified (this analysis does not require a called genotype and therefore exceeds the 12,610 overweight or obese participants in the primary analytic sample). Men under-classified more often than women (73.5% vs 51.6%; adjusted OR for women vs men 0.41, 95% CI 0.38–0.44, p < 10⁻¹²⁰). Excluding the 1,713 participants who described themselves as lean/fit (13.3% of this group, 88.0% of them men), under-classification fell to 58.4% and the male excess narrowed without disappearing (66.6% of men vs 49.7% of women; adjusted OR 0.52, 95% CI 0.48–0.56). Under- classification decreased with increasing BMI (adjusted OR 0.77 per 5 kg/m², p < 10⁻⁶⁴), so participants further into the obese range recognised their status more often. It also decreased with age, from approximately 66–68% below 40 years to 51.4% at 70 years and above (adjusted OR 0.89 per decade, 95% CI 0.87–0.92, p = 3 × 10⁻¹³), and the age gradient was steeper in men than in women (interaction p = 0.018). Estimates are mutually adjusted for age, sex and BMI as applicable. Across self-reported ethnic groups, under-classification ranged from 53.8% in Asian participants (n = 459) to 67.4% in participants of mixed ethnicity (n = 129).

Because BMI does not distinguish fat from lean mass, we repeated the comparison using waist-to-height ratio (WHtR ≥ 0.5) in the subset with waist measurements (n = 4,270). The sex difference persisted but was smaller: 40.7% of men and 21.7% of women with WHtR ≥ 0.5 did not perceive themselves as overweight or obese, compared with 56.4% and 26.3% of those overweight or obese by BMI in the same subset. Among participants overweight or obese by BMI who did not describe themselves as either, 68.6% of those in the overweight band had WHtR below 0.5, whereas in obese class I only 36.8% did; BMI may therefore overstate misperception in the overweight band, but not in the obese bands (Supplementary Table S6).

### 3.5 Genetic risk does not predict under-perception; recalled childhood obesity does

Overweight/obese self-perception increased with FTO risk genotype: 35.1% of low-risk (TT), 37.8% of moderate-risk (TA) and 39.9% of high-risk (AA) participants described themselves as overweight or obese (Figure 2A). Adjusted for age and sex, the high- versus low-risk difference was 4.7 percentage points (95% CI 2.7–6.7), and each risk allele added 2.4 percentage points. The estimate was essentially unchanged by adjustment for genotyping source, and it persisted after adjustment for self-reported ethnicity (+3.9 percentage points, 95% CI 1.2–6.6; high- versus low-risk contrast, n = 6,130, among the 11,571 participants who reported ethnicity), in logistic models (high versus low risk OR 1.22, 95% CI 1.12–1.33; per-allele OR 1.11) and in White participants alone (+3.7 percentage points, 95% CI 0.7–6.6, p = 0.015). The combined FTO+MC4R score showed a similar gradient, from 34.8% at a score of 0 to 43.9% at a score of 4 (+1.7 percentage points per risk allele, 95% CI 1.0–2.4, p = 4 × 10⁻⁶). MC4R alone was not associated with self-perception (+0.9 percentage points per allele, 95% CI −0.2 to +2.0, p = 0.11), so the smaller per-allele estimate for the score reflects dilution of the FTO signal rather than a contribution from MC4R (Supplementary Table S2).

**Figure 2.**
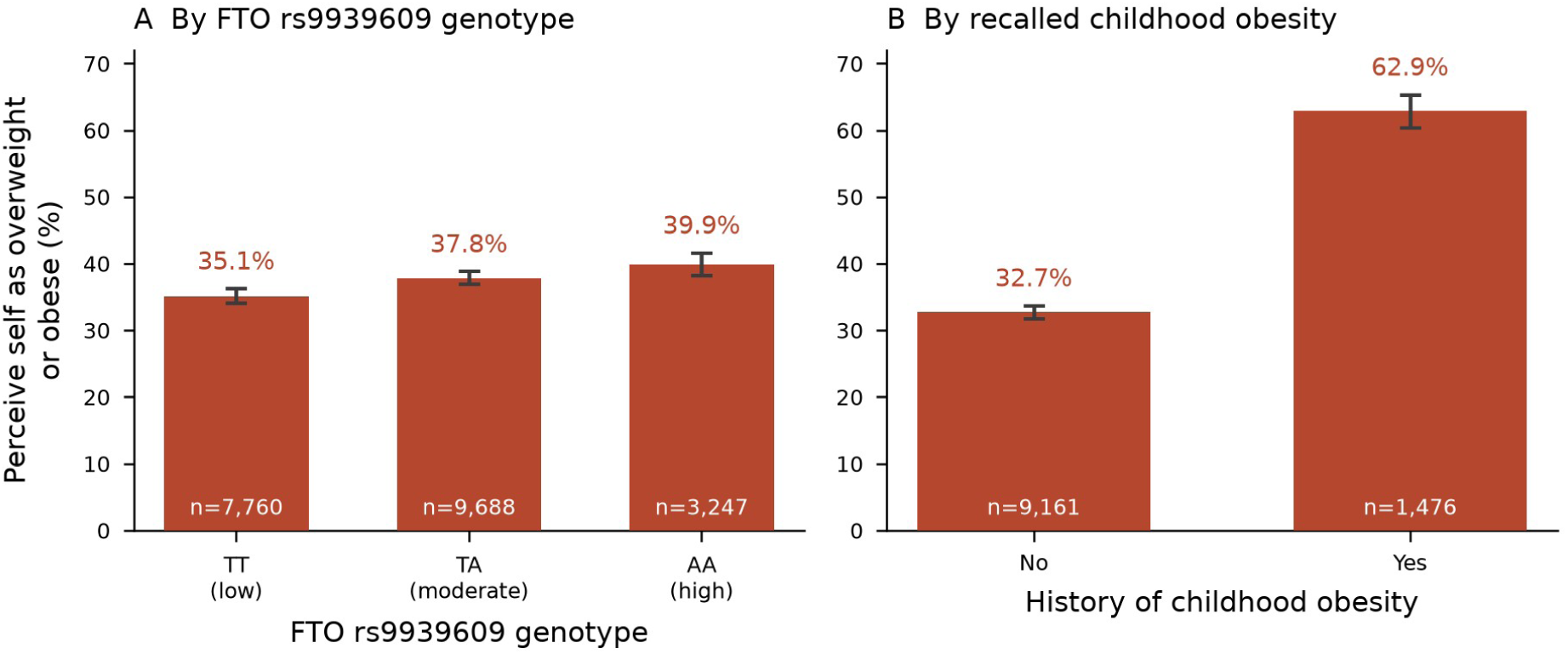
Overweight/obese self-perception by (A) FTO rs9939609 genotype (association sample, N = 20,695) and (B) recalled history of childhood obesity (n = 10,637; the association sample with the item answered, see Supplementary Table S3). Bars are unadjusted percentages; error bars are 95% Wilson confidence intervals; group sizes are shown within each bar.

These associations were fully explained by body size. Adjusted for BMI, the FTO high- versus low-risk difference fell to 0.6 percentage points (95% CI −1.0 to +2.2), and the score association was abolished (p = 0.25). To test directly whether genotype biases perception at a given body size, we modelled under- classification among participants with BMI ≥ 25 kg/m², adjusted for BMI, age and sex. Neither variant was associated with under-classification: FTO per-allele OR 0.97 (95% CI 0.92–1.03, p = 0.31); FTO high versus low risk OR 0.91 (95% CI 0.82–1.02, p = 0.10); MC4R per-allele OR 0.99 (95% CI 0.93–1.05, p = 0.71); FTO+MC4R score OR 0.98 (p = 0.33). The FTO estimate was null in men and women separately (sex interaction p = 0.63). The analysis had 80% power to detect per-allele effects larger than approximately 8% on the odds scale. At a given BMI, therefore, any effect of either variant on self- perception is small: the 95% confidence interval for the per-allele FTO estimate runs from 8% lower to 3% higher odds.

The per-allele association of FTO with self-perception was larger in women (+3.2 percentage points, 95% CI 1.8–4.6) than in men (+1.6, 95% CI 0.3–2.9), in parallel with its larger effect on BMI in women (Section 3.2), but the sex-by-genotype interaction was not significant (p = 0.12). An exploratory interaction between FTO genotype and the ordinal physical-activity score was also not significant (−0.26 percentage points per allele per activity level, p = 0.53; n = 20,691). Substituting WHtR for BMI in the waist subset (n = 4,171) gave the same pattern, although the subset is underpowered for FTO: the age- and sex-adjusted per-allele association was +1.75 percentage points (95% CI −0.36 to +3.86) and was removed by adjustment for WHtR (+0.11, 95% CI −1.64 to +1.85), as it was by adjustment for BMI (−0.68, 95% CI −2.28 to +0.91) (Supplementary Note S2).

In contrast to genotype, a recalled history of childhood obesity was associated with self-perception independently of current body size. Participants who reported childhood obesity described themselves as overweight or obese more often than those who did not (62.9% vs 32.7%; +31.5 percentage points adjusted for age and sex; Figure 2B), a difference 6.7 times the size of the high-versus-low genotype contrast (+4.7 percentage points). After adjustment for current BMI, age and sex, childhood obesity remained associated with a 10.0-percentage-point higher probability of overweight/obese self- perception (95% CI 7.4–12.5, p = 4 × 10⁻¹⁴; n = 10,797) and, among participants overweight or obese by BMI, with less under-classification (OR 0.64, 95% CI 0.56–0.74; n = 6,595).

An exploratory mediation analysis in the subset with recalled childhood obesity (n = 5,626) confirmed that the route from genotype to self-perception through childhood obesity also runs mainly through current body size: after adjustment for current BMI, the component of the genotype association passing through childhood obesity was +0.4 percentage points, 13% of the total. Supplementary Figure S1 shows the decomposition before current BMI is taken into account, in which the indirect component is +1.3 percentage points, approximately 40% of the total but with a confidence interval too wide to interpret.

### 3.6 Under-perception is associated with fewer weight-loss goals

Goals were recorded for 19,221 participants: get fit (46.0%), lose weight (25.6%), gain lean muscle (15.1%) and body maintenance (13.3%); 19,156 also had a self-perceived body type and a BMI category. Within every BMI category, participants who perceived themselves as overweight or obese set a weight- loss goal more often than those who did not (Figure 3A): 39.4% vs 10.7% at normal BMI, 42.2% vs 15.8% at overweight BMI and 49.4% vs 18.0% at obese BMI — gaps of 29, 26 and 31 percentage points within the same BMI category. Across the five perceived categories, the proportion setting a weight-loss goal rose from 2.9% (underweight) and 5.8% (lean/fit) to 18.7% (average), 43.8% (overweight) and 54.9% (obese).

**Figure 3.**
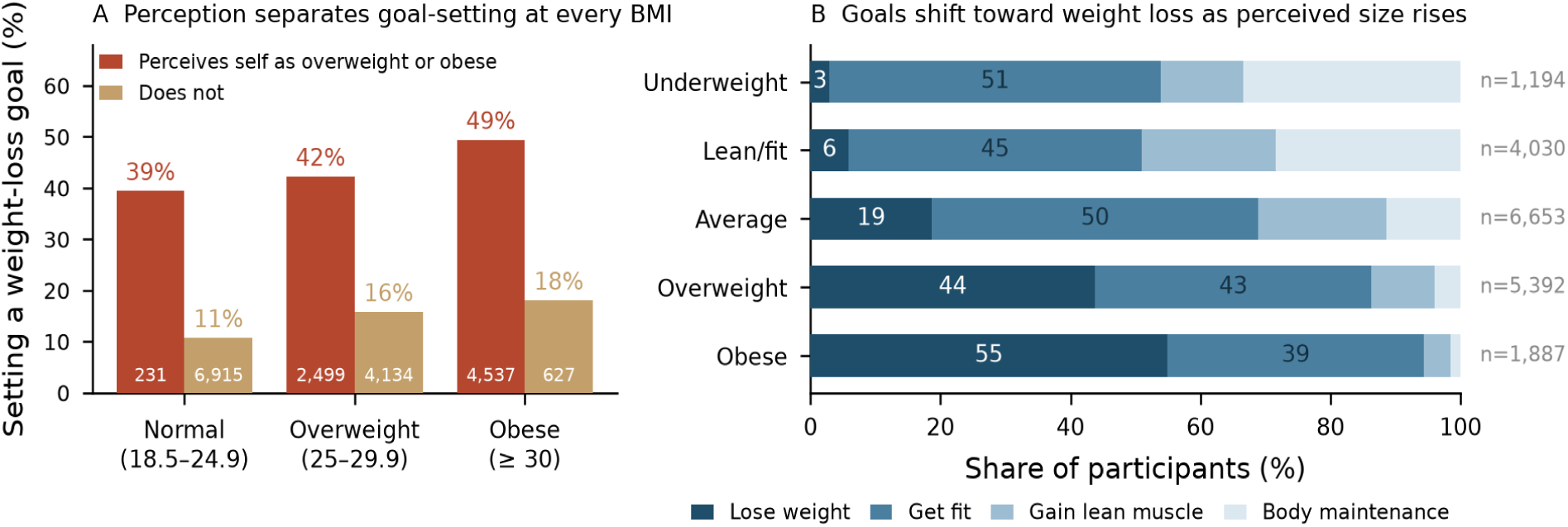
Weight-loss goals by self-perception and BMI category. (A) Participants setting a weight-loss goal, by BMI category and by whether they perceived themselves as overweight or obese (N = 18,943); cell counts are shown within each bar. Obesity classes I–III are combined because only 34 and 66 participants in classes II and III respectively did not perceive themselves as overweight or obese; the 213 participants with underweight BMI are omitted, only 12 of whom perceived themselves as overweight or obese. (B) Composition of the four weight-management goals across the five self-perceived body types, ordered from self-described obese at the top to underweight at the bottom (N = 19,156; row totals at right). Read from underweight to obese the weight-loss share rises monotonically (2.9% to 54.9%) and the body-maintenance share falls monotonically (33.4% to 1.5%), while the share choosing to get fit varies comparatively little (50.9% to 39.5%); segment labels are shown for the two largest goals only. Unadjusted percentages throughout.

Under-classification was associated with fewer weight-loss goals. Among the 11,797 participants who were overweight or obese by BMI and had recorded both a body type and a goal, 26.8% of those who under-classified set a weight-loss goal, against 47.5% of those whose perception matched their BMI category (OR 0.52, 95% CI 0.48–0.56, adjusted for BMI, age and sex). The association was present in men and women separately and in every age band.

With perception and BMI entered together and adjusted for age and sex, perceiving oneself as overweight or obese was associated with a 26.5-percentage-point higher probability of a weight-loss goal (95% CI 24.7 to 28.2), whereas each additional unit of BMI added 0.7 percentage points (95% CI 0.6 to 0.9). Adding perception to the model reduced the BMI coefficient from 2.0 to 0.7 percentage points per kg/m², a reduction of approximately 63%. Because a binary perception contrast and a per-unit BMI slope are not on a common scale, we also expressed both per standard deviation of the predictor (BMI 6.5 kg/m²; perception 0.49). Entered separately, the two were of similar strength: +15.8 percentage points per SD for perception (95% CI 15.2 to 16.4) and +12.7 for BMI (95% CI 12.0 to 13.4). Entered together, perception retained +12.9 percentage points per SD (95% CI 12.0 to 13.7) while BMI fell to +4.7 (95% CI 3.8 to 5.5). The difference between the two predictors therefore lies in how each survives adjustment for the other rather than in their separate predictive strength. This is not a mediation estimate: self-reported BMI is measured with error, which attenuates its coefficient, and perception may partly capture adiposity that BMI misses. A second adiposity measure, however, changed the perception estimate little. In the subset with waist measurements, the perception estimate was +24.1 percentage points adjusted for BMI, +23.4 adjusted for BMI and WHtR jointly, and +28.6 adjusted for WHtR alone. Logistic models gave the same pattern (perception OR 4.29, 95% CI 3.93–4.69; BMI OR 1.03 per kg/m²).

The perception–goal association was robust to model specification. In a multinomial model across all four goals, taking ‘get fit’ as the reference category, perceiving oneself as overweight or obese raised the odds of a weight-loss goal 3.34-fold (95% CI 3.04–3.66) and lowered the odds of gaining lean muscle (OR 0.57) and of body maintenance (OR 0.36), each relative to ‘get fit’. Treating perception as a five- level ordered variable gave +13.2 percentage points per category (95% CI 12.5 to 13.9), and adjustment for physical activity left the binary estimate at +23.4 percentage points. An unmeasured confounder would require a risk-ratio association of at least 5.43 with both perception and goal-setting to account for the association (E-value; lower confidence limit 5.06).

Because perception and goal were reported in the same questionnaire, part of their association could be definitional. Three observations argue against a purely definitional link. First, goals were substituted rather than relabelled (Figure 3B). From self-described underweight to obese, the share choosing to get fit varied little (50.9%, 45.1%, 50.2%, 42.5%, 39.5%), whereas the rise in weight-loss goals (2.9% to 54.9%) was matched by a monotonic fall in body maintenance (33.4% to 1.5%) and, from the ‘average’ category onwards, in gaining lean muscle (19.7% to 4.1%); a semantic restatement would not produce that particular exchange between goals. Second, recognition did not determine the goal: of the 7,279 participants who perceived themselves as overweight or obese, 46.7% set a weight-loss goal and 53.3% chose another, 78.3% of whom chose to get fit. Third, 12.7% of participants who did not perceive themselves as overweight or obese set a weight-loss goal, and broadening the outcome to any weight- or fitness-framed goal left a substantial association (+20.6 percentage points).

The size of the association depended on questionnaire completeness. It was +34.7 percentage points (95% CI 32.3 to 37.1) among participants who completed the extended questionnaire module and +14.2 (95% CI 11.9 to 16.6) among those who did not (difference +20.5, 95% CI 17.1 to 23.8, p = 7 × 10⁻³³). Module completion explains this difference; ancestry does not, and the larger estimate in White participants alone (+34.4 percentage points) follows from how many of them completed the module. Participants who did not complete the module also set weight-loss goals less often (16.7% vs 32.8%). That lower rate, together with the much sparser questionnaire records in this group, may reflect a less deliberate or partly default onboarding response rather than a different relationship between perception and intention. Because the outcome was also less common in this group, part of the difference on the percentage-point scale is expected from the lower base rate alone; on the odds scale the association was OR 5.19 (95% CI 4.64–5.82) in participants who completed the module and OR 3.11 (95% CI 2.67–3.63) in those who did not, a narrower difference than the percentage-point contrast implies. In either case, the pooled estimate should be read as an average over two groups whose data differ in completeness.

Genotype did not predict goal-setting beyond body size. FTO risk was weakly associated with a weight- loss goal when adjusted for age and sex (+1.2 percentage points per allele, 95% CI 0.3 to 2.0), but not after adjustment for BMI (+0.2, 95% CI −0.7 to 1.0), and the FTO+MC4R score was null after BMI adjustment (+0.0, 95% CI −0.6 to 0.7). Childhood obesity was associated with goal-setting at the same current BMI (+3.7 percentage points, 95% CI 1.0 to 6.4), but not once current self-perception was included (+0.2, 95% CI −2.4 to 2.7).

### 3.7 Men recognised overweight less often and, having recognised it, chose weight loss less often

The two steps compound in men. Among the 11,797 participants who were overweight or obese by BMI with both a body type and a goal recorded (6,533 men, 5,264 women), fewer men than women described themselves as overweight or obese at all: 47.0% against 75.4%, so 53.0% of men did not recognise their weight against 24.6% of women. The broader under-classification measure runs in the same direction (73.5% of men vs 51.6% of women; Section 3.4). Among those who did describe themselves as overweight or obese, men then set a weight-loss goal less often than women (42.0% vs 50.2%; adjusted OR for women vs men 1.38, 95% CI 1.26–1.52, p = 1 × 10⁻¹¹), choosing to gain lean muscle more often instead (12.5% vs 4.9%).

The net effect of the two steps is large. In the same sample, 25.5% of men set a weight-loss goal against 45.4% of women — a gap of 19.9 percentage points. Assigning men the female recognition rate while holding their own goal rates constant raises the male figure to 34.5%, so approximately 45% of the gap follows from the first step and the remainder from the second. This decomposition is descriptive and depends on the order in which the two steps are substituted, but it shows that neither step accounts for the difference on its own.

## 4 Discussion

Most adults who were overweight or obese by BMI placed themselves in a lighter category than their BMI indicated, and those who did so were about half as likely to set a weight-loss goal as participants of the same BMI who did not. Under-perception was concentrated in men, younger adults and those at lower BMI within the overweight and obese range. It was not explained by genetic risk of obesity: FTO rs9939609 raised BMI and the odds of childhood obesity, but at a given BMI neither it nor MC4R predicted how participants saw their weight. A recalled history of childhood obesity, by contrast, was associated with heavier self-perception at the same adult BMI. The rest of this section takes these strands in turn: who misperceived and why, what recognition predicts, what genotype and early history did and did not do, and what this means for digital weight-management services.

The extent and direction of misperception agree with previous reports that overweight and obese adults frequently perceive themselves as lighter than they are (Duncan et al., 2011; Gruszka et al., 2022; Shabu et al., 2024); under-classification outnumbered over-classification by more than twenty to one (44.8% vs 2.0% under the primary mapping). The ‘lean/fit’ response option describes fitness as well as size, so men may select it on the basis of muscularity. The sex difference was smaller when waist-to-height ratio rather than BMI was the reference, consistent with this explanation, although no direct measure of body composition was available and the excess was present on both measures. The cross-sectional design also cannot separate age from birth cohort. Younger cohorts, raised amid higher population adiposity, may have recalibrated their reference for normal weight upward (Robinson, 2017), which would produce the same gradient; we therefore describe it as an age–cohort gradient rather than as change within individuals.

Recognition tracked the weight-management goal participants set. At the same BMI, perceiving oneself as overweight or obese was associated with a 26.5-percentage-point higher probability of setting a weight-loss goal, and under-classification with approximately half the odds. Placed on a common scale, perception and BMI were of similar strength when entered separately (+15.8 and +12.7 percentage points per standard deviation), but when entered together perception retained +12.9 and BMI fell to +4.7: adding perception absorbs most of the association between measured size and the goal set, whereas adding measured size absorbs little of the association between perception and the goal. Error in self-reported BMI would push the estimates in the same direction, so the asymmetry may be overstated. Because perception and goal were reported in the same questionnaire, we do not claim that recognition causes intention; the two may be aspects of a single self-appraisal, and their association is partly definitional. Three observations nonetheless suggest that perception predicts which goal a person selects rather than simply restating it: goals were substituted rather than relabelled across perceived categories, most participants who recognised overweight chose a goal other than weight loss, and the association persisted when the outcome was broadened. The association was weaker among participants who did not complete the extended questionnaire module (+14.2 vs +34.7 percentage points), so the pooled estimate averages over two groups whose data differ in completeness.

In men, the two steps compound. Men were less likely to recognise overweight (among those overweight or obese by BMI, 47.0% of men described themselves as overweight or obese against 75.4% of women) and, having recognised it, less likely to set a weight-loss goal (42.0% vs 50.2%), choosing to gain lean muscle more often instead. The two steps together leave 25.5% of men who were overweight or obese by BMI setting a weight-loss goal, against 45.4% of women. The ‘lean/fit’ option and the smaller sex difference against waist-to-height ratio suggest that muscularity explains part of the first step, but not all of it, and it does not explain the second.

FTO risk was associated with heavier self-perception only because carriers are heavier. Adjustment for BMI abolished the association, and at a given BMI neither FTO, MC4R nor their combined score predicted misperception, with the FTO estimate bounded between OR 0.93 and 1.02. To our knowledge, this is the first explicit test of a perception-specific effect at the FTO locus. The result parallels FTO’s associations with type 2 diabetes and related metabolic traits, which are explained by its effect on BMI (Frayling et al., 2007; Freathy et al., 2008), and contrasts with its association with depression, which appears to persist, or even reverse, after BMI adjustment (Samaan et al., 2013). Because BMI adjustment does not remove every FTO association, the null observed here is informative. FTO has been linked to eating behaviour and affect beyond BMI (Harbron et al., 2014; Koike et al., 2018; Brown et al., 2026), but our results provide no evidence that these links extend to how carriers perceive their body size.

Two analytical choices qualify this conclusion. Restricting the analysis to BMI ≥ 25 kg/m² and adjusting for BMI both condition on a variable that FTO influences. If unmeasured factors affect both BMI and self- perception, such conditioning can bias the estimate in either direction (VanderWeele, 2016). Bias of this kind would need to be large to produce an estimate so tightly centred on the null, but it cannot be excluded, and the BMI-adjusted estimate answers a conditional question rather than providing an unbiased causal contrast.

At a given BMI, self-perception was associated with developmental history rather than with genotype. A recalled history of childhood obesity was associated with heavier self-perception, and with better recognition of overweight, at the same current BMI. This is consistent with prospective evidence that childhood adiposity predicts later body dissatisfaction (Blundell et al., 2024) and with a lasting effect of early body size on adult body image. Childhood obesity was more common among FTO risk-allele carriers and marked a heavier adult trajectory, but the share of the genotype–perception association that passed through it operated mainly through current BMI. This finding is the most exposed to bias. Childhood obesity was recalled retrospectively and reported in the same questionnaire as self- perception, recall of early-life body size is only moderately accurate (De Rubeis et al., 2019), and current self-image may shape recall. Its robustness to unmeasured confounding is accordingly modest: the E- value for the BMI-adjusted association is 1.91 (lower confidence limit 1.72), against 3.34 before BMI adjustment (Supplementary Note S1). Longitudinal data are required to separate a developmental effect from correlated recall.

These results have two implications for digital health services. First, genotype at these loci identifies people at higher risk of weight gain but not those who under-perceive their weight; current BMI and early weight history carry that information. Genetic risk reports may therefore be more useful when paired with objective feedback on current weight status than when presented alone. Second, services that rely on self-referral or self-set goals will under-reach the groups least likely to recognise their weight (men, younger adults and those at lower BMI within the overweight and obese range), because at the same BMI participants who under-classified were approximately half as likely to set a weight-loss goal. Whether presenting objective weight status, or framing feedback around the discrepancy between perceived and measured size, changes goal-setting remains untested. We observed stated intention at a single time point and did not analyse engagement or weight outcomes, which are themselves associated with digital engagement in weight-management services, including pharmacotherapy-supported programmes (Johnson et al., 2025).

The study also illustrates what DTC genetics cohorts can contribute to the study of health behaviour. They include people before, and independently of, any clinical encounter, and they link genotype to perception, developmental history and stated intention in the same individuals at scale. People who have not identified a weight concern and would not present for care are rarely represented in clinical datasets, yet they are the population in which the perception effects reported here matter most. Using such data to personalise body-image or health communication is a hypothesis to test prospectively, not an established application.

Several limitations apply. The design is cross-sectional: mediation is estimated statistically and does not establish causation, and temporal ordering is not observed. Height and weight were self-reported, and self-report typically underestimates weight (Connor Gorber et al., 2007); because the primary outcome is subjective perception, this matters less than it would for a pure misperception analysis, but residual bias remains. Recalled childhood obesity is subject to the recall limitations described above and was available for only about half the cohort, a subset defined by completion of an extended questionnaire module rather than at random (Supplementary Table S1); the mediation analysis therefore rests on a non-random subsample, and we treat it as exploratory. The concordance estimate depends on how the ambiguous ‘average’ label is mapped (53–68%). Genotypes departed mildly from Hardy–Weinberg equilibrium, which may reflect population structure in a multi-ancestry cohort of mixed genotyping provenance or a genotyping artefact in the array subset and cannot be resolved with these data; ancestry was captured by self-reported ethnicity rather than genetic principal components, and ethnicity was missing for 44.1% of participants, so ancestry-stratified analysis is warranted. The cohort is a self-selected sample of paying customers, more health-motivated and demographically skewed than the general population, and self-perceived body type was measured with a single non-validated item.

Primary estimates are adjusted only for age and sex, and confounders of the relation between childhood obesity and self-perception, such as socioeconomic position, were unmeasured. MC4R is a weak marker of obesity risk in this cohort: its own association with BMI did not reach significance (+0.14 kg/m² per allele, p = 0.07), so its null results are less informative than FTO’s. Two variants explain only a small fraction of BMI variance, so these findings do not address polygenic obesity risk more broadly. Finally, perception and goal were reported in the same questionnaire, so their association is exposed to shared- method variance. These constraints limit causal inference but do not alter the central observation: adults’ stated weight-management intention follows how they perceive their weight more closely than it follows their measured size or the FTO and MC4R variants examined here.

In conclusion, around two in five adults who were overweight or obese by BMI did not describe themselves as such, most often men, younger adults and those at lower BMI within the overweight and obese range, and at the same BMI those who placed themselves in a lighter category than their BMI were about half as likely to set a weight-loss goal. The obesity risk allele at FTO was associated with higher body size in childhood and adulthood but, at a given BMI, not with how adults saw their weight; a recalled history of childhood obesity was, a finding that requires longitudinal confirmation. Genetic risk therefore identifies who is likely to be heavier but not who will recognise it. Self-perception is a candidate target for intervention, and whether feedback on objective weight status can recalibrate it, and thereby change the goals people set, is a question for prospective studies.

## Data Availability Statement

Individual-level consumer genetic and questionnaire data cannot be shared openly: the consent recorded at registration and the company’s privacy commitments do not extend to open release of individual records. Aggregate data sufficient to reproduce every result reported here will be deposited alongside the analysis code — the full self-perceived body type × BMI category contingency table underlying Figure 1, the goal × perceived body type × BMI category counts underlying Figure 3, and the summary tables underlying Table 1 and Supplementary Tables S1–S6. Requests for further aggregate output may be addressed to the corresponding author and will be considered subject to FitnessGenes data-governance approval. Analysis code (data preparation and statistical models) will be deposited in a public repository (Zenodo) with a DOI issued at publication.

## Ethics Statement

The study was reviewed by the FitnessGenes internal Ethics Committee (reference FGEC/2026/012), which confirmed valid opt-in research consent, compliance with UK GDPR, and anonymisation of the dataset. The study was conducted in accordance with the Declaration of Helsinki and the local legislation and institutional requirements. Participants provided separate, opt-in electronic informed consent for the research use of their data at registration, before genetic testing, and could withdraw consent at any time. The research team had no access to personal identifying information.

## Author Contributions

OMM: Conceptualization, Methodology, Formal analysis, Writing – original draft. NRW: Data curation, Software, Writing – review & editing. SJG: Supervision, Writing – review & editing. All authors contributed to the article and approved the submitted version.

## Generative AI Statement

The author(s) declare that Generative AI was used in the creation of this manuscript. The authors used Claude (Anthropic) for statistical verification, re-analysis and language editing. Estimates added or revised during that process were recomputed from the source data before inclusion; the authors checked and approved all content and take full responsibility for the manuscript.

## Data Availability

All data produced in the present study are available upon reasonable request to the authors

## Acknowledgments

We thank the FitnessGenes customers who consented to the research use of their data, and the FitnessGenes team for data management and technical support.

## Supplementary Material

The following items are provided as Supplementary Material: Supplementary Tables S1–S6, Supplementary Figure S1 and Supplementary Notes S1 and S2.

---

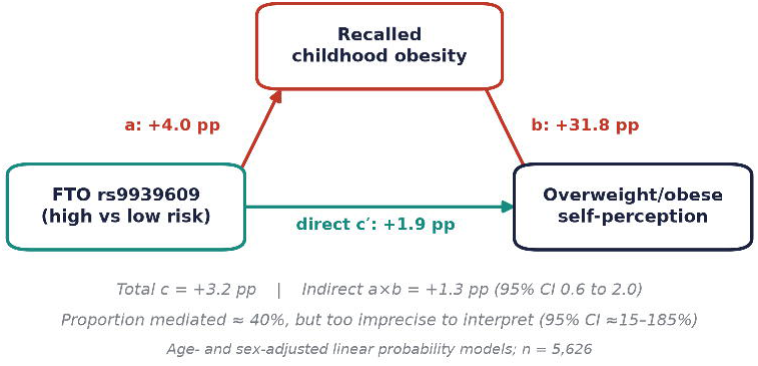

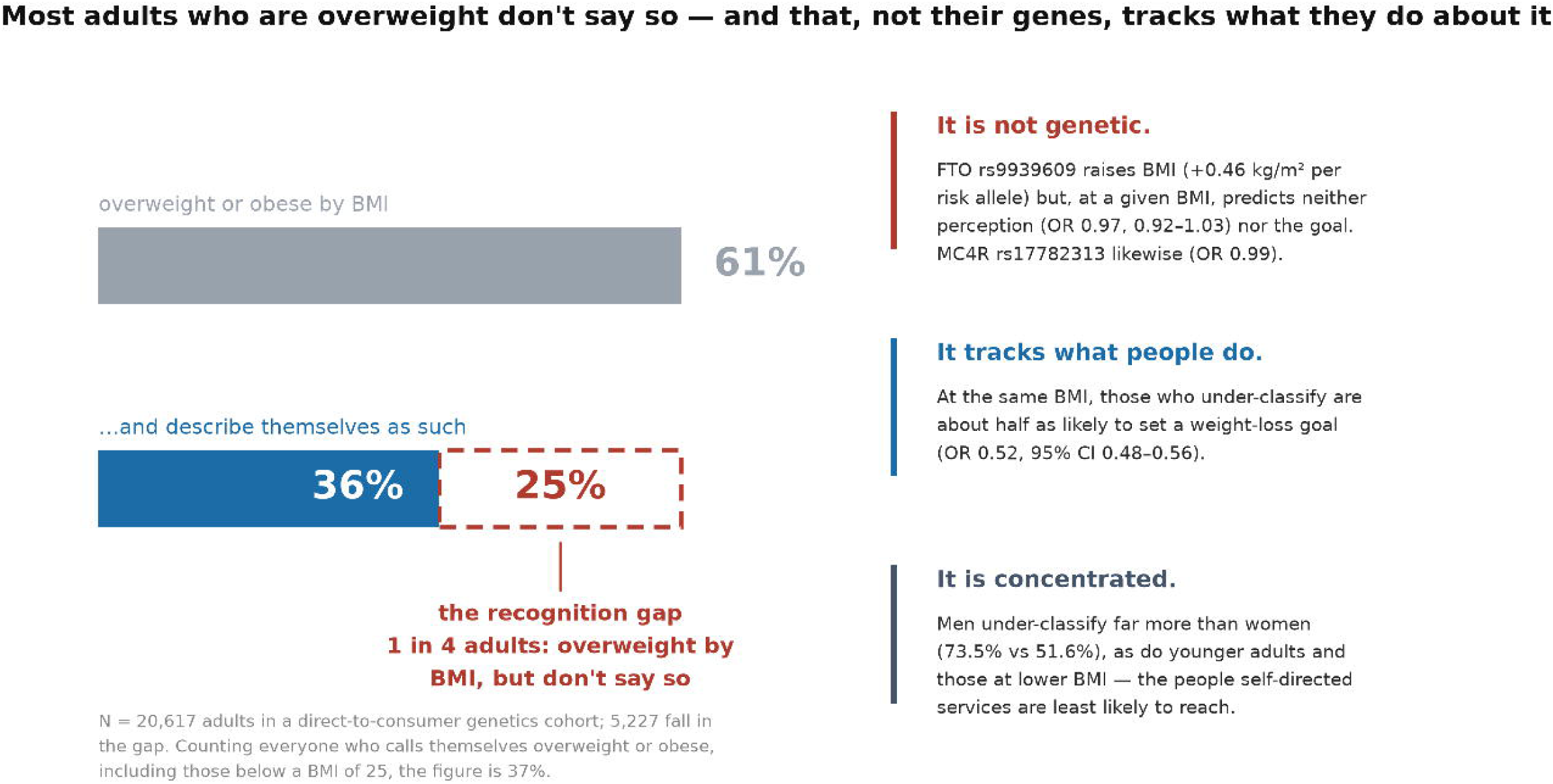

