## Supplementary for "Under-perception of overweight, not genetic risk, tracks weight-loss goal-setting in a direct-to-consumer genetics cohort"

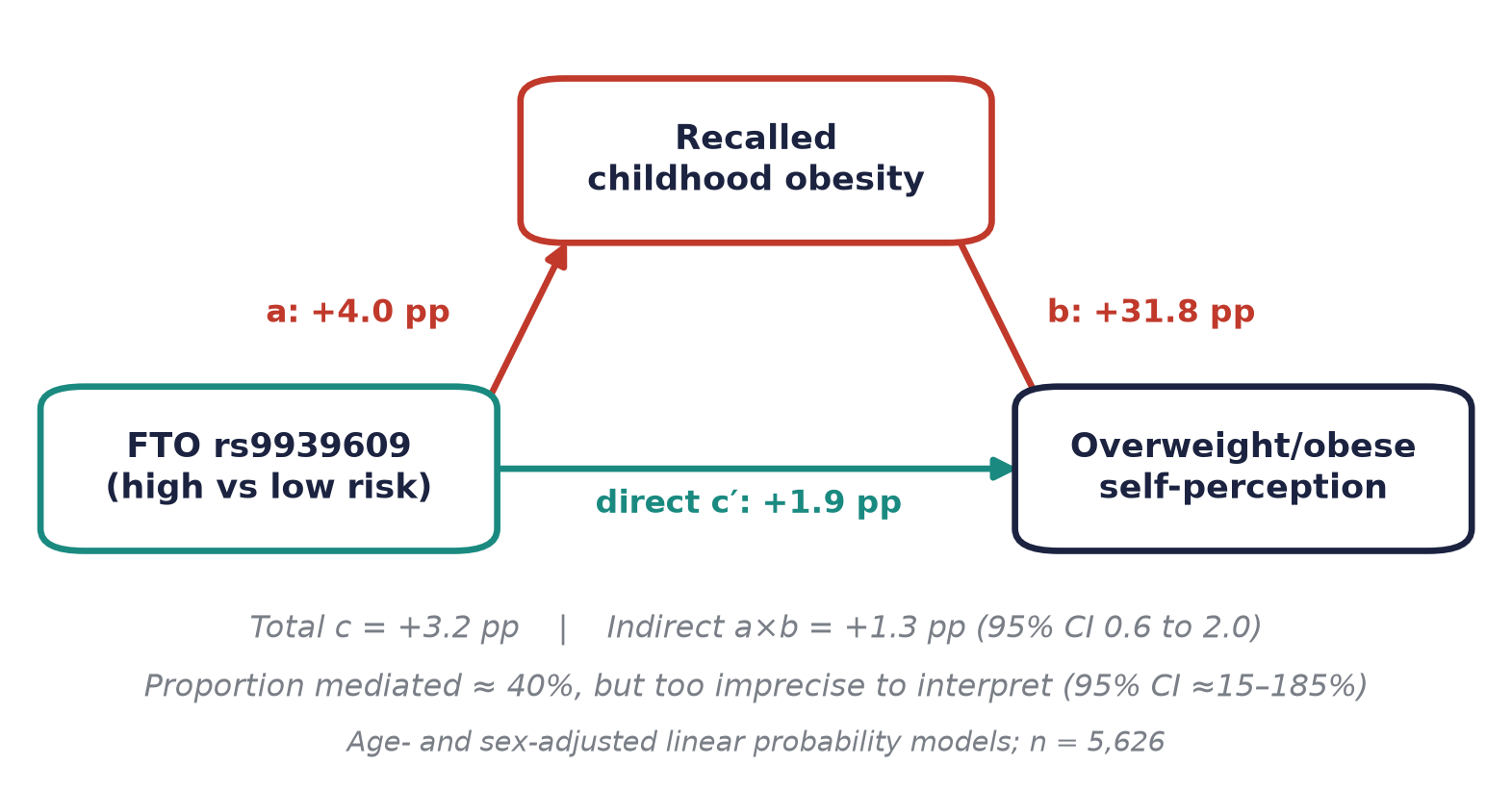

Supplementary Figure S1. Age- and sex-adjusted mediation of the FTO → self-perception association by recalled childhood obesity (N = 5,626). Path a, effect of high- vs low-risk genotype on recalled childhood obesity; b, effect of recalled childhood obesity on self-perception, adjusted for genotype; c′, direct effect; c, total effect. All paths from linear-probability models; pp, percentage points.

Supplementary Table S1. Characteristics of participants with and without recalled childhood-obesity data (primary analytic sample, N = 20,617). Standardised differences are reported alongside p-values because at this sample size trivial differences reach significance; |d| < 0.1 indicates good balance. The two groups are closely balanced on body size and on self-perceived overweight or obesity, but differ markedly in questionnaire completeness: the childhood-obesity item belongs to an extended module that also carried the waist, diabetes, family-history and emotional-eating items.

| **Characteristic** | **With childhood-obesity data** | **Without** | **Standardised difference** | **p** |
| --- | --- | --- | --- | --- |
| n | 10,609 | 10,008 |  |  |
| Age, mean (SD), years | 46.2 (15.5) | 44.3 (22.2) | +0.096 | 7.9e-12 |
| Female, % | 46.4 | 50.0 | -0.072 | 3.0e-07 |
| BMI, mean (SD), kg/m² | 27.5 (6.2) | 27.9 (6.9) | -0.072 | 2.9e-07 |
| Overweight or obese by BMI, % | 61.0 | 61.4 | -0.008 | 0.562 |
| BMI category, % (normal / overweight / obese I / obese II / obese III / underweight) | 37.9 / 36.1 / 15.2 / 5.6 / 4.1 / 1.1 | 37.3 / 33.4 / 15.5 / 6.9 / 5.6 / 1.3 | — | 1.9e-10 |
| Perceives self as overweight or obese, % | 36.9 | 37.4 | -0.009 | 0.515 |
| Self-perceived body type, % (underweight / lean-fit / average / overweight / obese) | 8.3 / 22.6 / 32.2 / 27.6 / 9.4 | 5.0 / 20.8 / 36.9 / 27.9 / 9.4 | — | 2.3e-26 |
| FTO rs9939609, % (TT / TA / AA) | 37.2 / 47.1 / 15.7 | 37.9 / 46.5 / 15.6 | — | 0.572 |
| FTO risk alleles, mean (SD) | 0.79 (0.70) | 0.78 (0.70) | +0.011 | 0.411 |
| Genotyping source, % (array / uploaded) | 63.8 / 36.2 | 72.2 / 27.8 | — | 2.0e-37 |
| Ethnicity reported, % | 98.8 | 10.5 | +3.849 | 0.0e+00 |
| Weight-loss goal, % | 32.3 | 14.0 | +0.443 | 2.0e-209 |
| Extended-module items present (waist, diabetes, family history, emotional eating), mean of 4 | 2.69 | 0.02 | +4.325 | 0.0e+00 |

Supplementary Table S2. FTO rs9939609, MC4R rs17782313 and two-variant scores across every key outcome. MC4R is reported on its own because the two variants act through partly different biology. Its own BMI association in this cohort is weak, so a matching null is supportive rather than a true internal replication. The weighted score uses per-allele BMI effects estimated in this cohort (FTO +0.46, MC4R +0.14 kg/m²); external weights can be substituted. All models adjust for age and sex; the under-classification models additionally adjust for BMI and are restricted to BMI ≥ 25.

| **Exposure** | **BMI (kg/m2)** | **Self-perceived overweight/obese (pp)** | **Under-classification, BMI>=25 (adj BMI)** | **Recalled childhood obesity** | **Weight-loss goal (pp)** | **Weight-loss goal, BMI-adjusted (pp)** |
| --- | --- | --- | --- | --- | --- | --- |
| FTO rs9939609 per allele | +0.46 kg/m2 (95% CI +0.33 to +0.59), p=6.1e-12 | +2.43 pp (95% CI +1.49 to +3.37), p=4.2e-07 | OR 0.97 (95% CI 0.92-1.03), p=0.31 | OR 1.18 (95% CI 1.09-1.28), p=3.4e-05 | +1.16 pp (95% CI +0.27 to +2.05), p=0.011 | +0.18 pp (95% CI -0.68 to +1.03), p=0.69 |
| MC4R rs17782313 per allele | +0.14 kg/m2 (95% CI -0.01 to +0.29), p=0.071 | +0.88 pp (95% CI -0.20 to +1.97), p=0.11 | OR 0.99 (95% CI 0.93-1.05), p=0.71 | OR 1.16 (95% CI 1.06-1.27), p=0.0014 | +0.15 pp (95% CI -0.87 to +1.17), p=0.78 | -0.18 pp (95% CI -1.16 to +0.80), p=0.73 |
| Unweighted 2-variant score | +0.31 kg/m2 (95% CI +0.21 to +0.41), p=2.2e-09 | +1.68 pp (95% CI +0.97 to +2.40), p=3.9e-06 | OR 0.98 (95% CI 0.94-1.02), p=0.33 | OR 1.17 (95% CI 1.10-1.24), p=3.5e-07 | +0.70 pp (95% CI +0.02 to +1.37), p=0.043 | +0.02 pp (95% CI -0.63 to +0.67), p=0.95 |
| BMI-weighted 2-variant score | +0.95 kg/m2 (95% CI +0.67 to +1.23), p=2.6e-11 | +5.11 pp (95% CI +3.11 to +7.11), p=5.7e-07 | OR 0.94 (95% CI 0.84-1.06), p=0.3 | OR 1.50 (95% CI 1.26-1.77), p=2.7e-06 | +2.37 pp (95% CI +0.48 to +4.27), p=0.014 | +0.30 pp (95% CI -1.53 to +2.12), p=0.75 |

Supplementary Table S3. Analytic samples used in the paper, with inclusion criteria and sizes. Section numbers refer to the main text. Several samples differ by one or two participants because individual models additionally require a covariate (age, sex or BMI); where this occurs both sizes are given.

| **Sample** | **n** | **Inclusion** |
| --- | --- | --- |
| Records in the de-identified extract | 21,293 | All rows supplied |
| Association sample | 20,695 | Non-missing FTO rs9939609 risk group and self-perceived body type |
| Primary descriptive sample (Table 1, Section 3.1) | 20,617 | Association sample additionally requiring a BMI category (78 otherwise-eligible participants lacked one) |
| High- vs low-risk contrast | 11,007 | Association sample restricted to TT or AA genotypes |
| Overweight or obese by BMI (Section 3.4) | 12,840 | BMI category overweight or obese with self-perceived body type; genotype not required |
| Goal-setting sample (Section 3.6) | 19,156 | Non-missing goal, self-perceived body type and BMI category |
| Overweight or obese within the goal-setting sample | 11,797 | Goal-setting sample with BMI category overweight or obese |
| Recalled childhood obesity, association sample (Figure 2B) | 10,637 | Association sample with the childhood-obesity item answered |
| Recalled childhood obesity, descriptive sample | 10,609 | Primary descriptive sample with the childhood-obesity item answered |
| Recalled childhood obesity, perception and BMI present | 10,798 | Perception and BMI category present and the childhood-obesity item answered; genotype not required. Models additionally requiring age and sex (Section 3.5) use 10,797, one participant fewer |
| Mediation subset (Section 3.5) | 5,626 | High- vs low-risk contrast with the childhood-obesity item answered and age and sex present |
| Ethnicity-adjusted contrast (Section 3.5) | 6,130 | High- vs low-risk contrast among participants reporting ethnicity |
| Waist subset, WHtR derivable (Section 3.2) | 4,271 | Waist circumference recorded and height available; recorded only within the extended questionnaire module. This is the Section 3.2 denominator |
| Waist subset with perception and BMI category (Section 3.4) | 4,270 | Waist subset additionally requiring self-perceived body type and a BMI category; genotype not required |
| Waist subset with a called genotype (Table S5) | 4,170 / 4,171 | Waist subset additionally requiring a called FTO genotype, age and sex; 4,171 for WHtR outcomes and 4,170 where BMI is also required |
| Central adiposity subset (Section 3.4) | 1,867 | Waist subset with WHtR >= 0.5, perception, BMI category and sex present |
| Implausible ages (reported, not excluded) | 32 | Age <18 or >100 y, consistent with placeholder birth dates; retained in primary analyses, see Section 2.1 |

Supplementary Table S4. Genotype quality control by source. rs9939609 is an A/T variant and therefore strand-ambiguous; its A-allele frequency is 0.391 in both the array and the uploaded subsets, whereas a systematic strand flip in the uploaded data would give approximately 0.61, so these data show no evidence of strand misalignment. Hardy–Weinberg deviation is confined to the array subset for both variants.

| **Variant** | **Source** | **n_called** | **Call_rate** | **Genotypes** | **Minor_allele_freq** | **HWE_p** |
| --- | --- | --- | --- | --- | --- | --- |
| FTO rs9939609 (A/T; strand-ambiguous) | Array | 14,265 | 99.9% | TT 37.6 / TA 46.6 / AA 15.8 | A = 0.391 | 0.011 |
| FTO rs9939609 (A/T; strand-ambiguous) | Uploaded | 6,664 | 95.1% | TT 37.4 / TA 47.1 / AA 15.5 | A = 0.391 | 0.363 |
| FTO rs9939609 (A/T; strand-ambiguous) | All | 20,929 | 98.3% | TT 37.5 / TA 46.8 / AA 15.7 | A = 0.391 | 0.009 |
| MC4R rs17782313 (A/G) | Array | 14,256 | 99.8% | AA 57.6 / AG 36.1 / GG 6.3 | G = 0.243 | 0.022 |
| MC4R rs17782313 (A/G) | Uploaded | 6,199 | 88.5% | AA 59.0 / AG 35.2 / GG 5.7 | G = 0.233 | 0.242 |
| MC4R rs17782313 (A/G) | All | 20,455 | 96.1% | AA 58.0 / AG 35.9 / GG 6.1 | G = 0.240 | 0.01 |

Supplementary Table S5. Genotype associations with waist-to-height ratio (WHtR) in the subset with waist measurements. Estimates are per risk allele from linear models (continuous WHtR, BMI in kg/m²) or logistic models (WHtR ≥ 0.5, odds ratio), with HC3 standard errors. “BMI within the waist subset” is reported to show that the subset is not atypical for genotype–adiposity associations. Because every model here requires a called genotype, the denominator is 4,171 (4,170 where BMI is also required) rather than the 4,271 and 4,270 used in main-text Sections 3.2 and 3.4, which do not require one (Supplementary Table S3). Denominators follow the genotype-call rule in Methods 2.3, under which the 15 ambiguous calls are treated as missing: 4,171 for FTO, 4,029 for MC4R and 4,022 for the two-variant score, each one lower where BMI is also required because one participant in each frame has no BMI value. Retaining the ambiguous calls would instead give 4,176, 4,029 and 4,026. At n = 4,171 the smallest per-allele effect on WHtR detectable with 80% power is 0.0057 units, so the unadjusted WHtR estimate is close to the limit of detection.

| Outcome | Exposure | Adjust | n | Estimate | LCL | UCL | p |
| --- | --- | --- | --- | --- | --- | --- | --- |
| BMI within the waist subset (kg/m2) | FTO | age+sex | 4,170 | 0.4764 | 0.2 | 0.7528 | 0.000729 |
| WHtR (continuous) | FTO | age+sex | 4,171 | 0.0056 | 0.0015 | 0.0097 | 0.00781 |
| WHtR (continuous) | FTO | age+sex+BMI | 4,170 | 0.0007 | -0.0021 | 0.0035 | 0.641 |
| WHtR >= 0.5 (OR) | FTO | age+sex | 4,171 | 1.1114 | 1.0173 | 1.2142 | 0.0193 |
| WHtR >= 0.5 (OR) | FTO | age+sex+BMI | 4,170 | 1.0006 | 0.8936 | 1.1204 | 0.992 |
| BMI within the waist subset (kg/m2) | MC4R | age+sex | 4,028 | 0.0841 | -0.2308 | 0.3989 | 0.601 |
| WHtR (continuous) | MC4R | age+sex | 4,029 | 0.0022 | -0.0026 | 0.0069 | 0.367 |
| WHtR (continuous) | MC4R | age+sex+BMI | 4,028 | 0.0013 | -0.002 | 0.0047 | 0.444 |
| WHtR >= 0.5 (OR) | MC4R | age+sex | 4,029 | 1.0597 | 0.9553 | 1.1756 | 0.273 |
| WHtR >= 0.5 (OR) | MC4R | age+sex+BMI | 4,028 | 1.0687 | 0.9375 | 1.2182 | 0.32 |
| BMI within the waist subset (kg/m2) | SCORE | age+sex | 4,021 | 0.2876 | 0.0777 | 0.4976 | 0.00725 |
| WHtR (continuous) | SCORE | age+sex | 4,022 | 0.0039 | 0.0008 | 0.0071 | 0.0152 |
| WHtR (continuous) | SCORE | age+sex+BMI | 4,021 | 0.001 | -0.0013 | 0.0032 | 0.396 |
| WHtR >= 0.5 (OR) | SCORE | age+sex | 4,022 | 1.0824 | 1.0111 | 1.1588 | 0.0227 |
| WHtR >= 0.5 (OR) | SCORE | age+sex+BMI | 4,021 | 1.0295 | 0.9438 | 1.123 | 0.512 |

Supplementary Table S6. Central adiposity among participants who did not recognise overweight, in the subset with waist measurements. Among participants overweight or obese by BMI, those who did not describe themselves as overweight or obese were largely not centrally adipose, and this was concentrated in the overweight band, where BMI and waist-to-height ratio diverge. Two limits apply: the extract contains no measure of fat or lean mass, so a low WHtR at raised BMI is consistent with muscularity but does not establish it — a favourable fat distribution would look the same; and physical activity did not distinguish the groups (70.3% of non-recognisers with WHtR < 0.5 were active or very active, against 67.5% of those with WHtR ≥ 0.5), so the pattern does not simply mark a more athletic subgroup. Under-recognition persists in the obese bands, where it carries the clearest clinical implication. Two denominators differ from the main text by design and are not discrepancies: 42.0% is the share not describing themselves as overweight or obese within the waist subset, against 41.5% among all participants overweight or obese by BMI (Section 2.3); and 64.2% is the share with WHtR < 0.5 among all non-recognisers in the waist subset, against 68.6% among those in the overweight band alone (Section 3.4).

| Analysis | Group | n | Metric | Value |
| --- | --- | --- | --- | --- |
| BMI>=25 with waist data | all | 2,761 | does not perceive overweight (%) | 42.0 |
| BMI>=25 with waist data | does not perceive overweight | 1,161 | WHtR < 0.5 (%) | 64.2 |
| BMI>=25 with waist data | does not perceive overweight | 1,161 | mean WHtR | 0.49 |
| BMI>=25 with waist data | does not perceive overweight | 1,161 | mean BMI | 27.75 |
| BMI>=25 with waist data | does not perceive overweight | 1,161 | active or very active (%) | 69.3 |
| BMI>=25 with waist data | perceives overweight | 1,600 | WHtR < 0.5 (%) | 19.6 |
| BMI>=25 with waist data | perceives overweight | 1,600 | mean WHtR | 0.571 |
| BMI>=25 with waist data | perceives overweight | 1,600 | mean BMI | 33.43 |
| BMI>=25 with waist data | perceives overweight | 1,600 | active or very active (%) | 34.2 |
| non-perceivers by BMI band | overweight 25-29.9 | 997 | WHtR < 0.5 (%) | 68.6 |
| non-perceivers by BMI band | obese I 30-34.9 | 152 | WHtR < 0.5 (%) | 36.8 |
| non-perceivers by BMI band | obese II+ >=35 | 12 | WHtR < 0.5 (%) | 41.7 |
| reference comparison | BMI >= 25 | 2,761 | does not perceive overweight (%) | 42.0 |
| reference comparison | WHtR >= 0.5 | 1,867 | does not perceive overweight (%) | 30.5 |

Supplementary Note S1. Sensitivity of the mediator–outcome association. For the age- and sex-adjusted mediator–outcome association (risk ratio 1.96), an E-value of 3.34 (lower confidence limit 3.15) indicates that an unmeasured mediator–outcome confounder would need a risk-ratio association of at least that magnitude with both childhood obesity and self-perception to explain the association away; for the BMI-adjusted association (risk ratio 1.30) the corresponding E-value is 1.91 (lower confidence limit 1.72). The indirect effect was estimated with 3,000 bootstrap resamples (percentile 95% confidence intervals).

Supplementary Note S2. Substituting waist-to-height ratio for BMI in the FTO–self-perception models. Section 3.5 tests whether FTO rs9939609 is associated with overweight/obese self-perception once body size is accounted for, using BMI. Because BMI does not distinguish fat from lean mass, the same models were refitted in the subset with waist measurements (n = 4,171; 2,229 for the high- versus low-risk contrast), substituting WHtR for BMI. Linear-probability models with HC3 standard errors, adjusted for age and sex, gave a per-allele association with self-perception of +1.75 percentage points (95% CI −0.36 to +3.86, p = 0.11) before adjustment for adiposity, +0.11 percentage points (95% CI −1.64 to +1.85, p = 0.90) after adjustment for WHtR, and −0.68 percentage points (95% CI −2.28 to +0.91, p = 0.40) after adjustment for BMI. The high- versus low-risk contrast behaved the same way: +3.98 percentage points (95% CI −0.47 to +8.43) adjusted for age and sex, −0.38 (95% CI −4.08 to +3.32) with WHtR added and −1.29 (95% CI −4.69 to +2.10) with BMI added. The subset is underpowered for FTO — the unadjusted estimate does not reach significance, unlike the +2.43 percentage points in the full sample — so this analysis cannot confirm the association, only show that whichever measure of adiposity is used, adjusting for it removes the estimate. The conclusion of Section 3.5 does not therefore depend on BMI being the adiposity measure.
